# Seroprevalence of the SARS-CoV-2 infection in health workers of the Sanitary Region VIII, at province of Buenos Aires

**DOI:** 10.1101/2020.09.07.20189050

**Authors:** Andrea P. Silva, M. Fernanda Aguirre, Christian Ballejo, M. Jimena Marro, Andrea V. Gamarnik, Gastón Vargas, Marina Pifano, Teresa B. Varela, Enio García, Alicia EB Lawrynowicz, Osvaldo C. Uez, Irene Pagano

## Abstract

**Introduction:** The aim of this study was to estimate the seroprevalence of the SARS-CoV-2 infection in health workers of the Sanitary Region VIII, at province of Buenos Aires during June 2020.

**Methods:** A cross-sectional design was used. A probabilistic sampling by two-stage conglomerates was carried out. Data were collected from a self-administered questionnaire and a blood sample for antibody identification. The COVIDAR IgG and IgM test were used. RESULTS: 738 health workers were included; the overall response rate was 73.80%. 71.83% of that were women; age showed a normal distribution. Nurses and doctors accounted for more than half of the staff. 75.86% of people claimed to always use Personal Protective Equipment. 5.61% of people had close contact with a confirmed case of COVID-19. 4.60% of people had previously had a nasopharyngeal swab with a negative result. Five workers had positive IgG for SARS-CoV-2 (four women and one man) with negative IgM. The mean age of the cases was 35 years old; two of them were asymptomatic; neither of them had a swab sample taken. The overall seroprevalence was 0.75%, with no significant differences between strata.

**Discussion:** The seroprevalence found was low; indicating a large proportion of workers was susceptible to infection. We stress the need to complement passive epidemiological surveillance strategies with serological monitoring in health workers.

## Introduction

At the end of 2019, the world witnessed the emergence of a new disease. A hospital from Wuhan, Hubei province, alerted the Chinese government about the occurrence of a number of strange cases of severe pneumonia which were reported to the World Health Organization (WHO). The pathogen involved was identified as a novel outbreak of coronavirus 2019 (SARS-CoV-2). On January 30, the WHO declared the situation as a public health emergency of international concern and, on March 11, it was characterized as a pandemic^1^.

COVID-19 pandemic, originated from infection of SARS-CoV-2, is significantly spreading around the world. Globally, on August 10, 2020, 19,718,030 cumulative cases and 728,013 deaths have been registered^2^. In Argentina, there were 253,868 confirmed cases and 4,785 deaths^3^.

In this context, the situation of health care workers represents a topic of interest because they are in the first line of response and their exposure to the virus is high as well as their potential to spread the infection in the community. This type of workers faces unprecedented occupational risks of morbidity and mortality^4–7^ which makes the timely implementation of protective measures imperative for this group^8–10^.

Health care workers are all those who provide health care services or whose job is to provide care services and who directly cooperate in assistance positions. This includes workers who are in charge of direct sick patient assistance (medical doctors, nurses, laboratory technicians, dentists, administrative workers, etc.) as well as those who are in charge of food, cleaning, maintenance and security services^9^. As of June 29, 2020, 9% (N = 2,761) of confirmed cases of SARS-CoV-2 infection in Buenos Aires province involved health care workers^11^.

The vast majority of the information available during the pandemic comes from passive epidemiological surveillance systems based on confirmation of suspected cases who have developed symptoms compatible with COVID-19. In this regard, the proportion of symptomatic infected individuals is mainly determined by RT-PCR from samples taken in nasopharyngeal and oropharyngeal swabs.

The strategies based on serological tests that measure antibody response on the population have been widely implemented in epidemiological studies^12^. Antibodies against the virus develop days or weeks after the infection so their presence accounts for the previous exposure to the virus and to the development of an immune response, regardless of the presence of symptoms. In this regard, the surveillance of antibody seropositivity as a public health strategy would allow us to reach to conclusions about the scope of the infection in a given population and the amount of asymptomatic individuals, and also, to detect cases that may have not been identified with passive surveillance strategies^13^. While many aspects regarding the duration and quality of the immune response to SARS-CoV-2 still need to be addressed, and considering that antibodies are markers of partial or total immunity, this type of studies could provide information as to which percentage of the population is susceptible to the virus^14^.

In harmony with the WHO recommendations regarding the implementation of early response to the pandemic by means serological studies^15^, Buenos Aires province set up an active epidemiological surveillance strategy which includes the serological monitoring of health care workers. Within this framework, a seroprevalence study which started with a pilot test in public hospitals of the Sanitary Region (RS, by its acronym in Spanish) VIII of the province was carried out.

The purpose was to estimate the SARS-CoV-2 infection seroprevalence on health care workers of the RS VIII public subsector in Buenos Aires province during June, 2020.

## Methods

A cross-sectional study was used. The target population was health care workers from public facilities from the RS VIII from Buenos Aires province during June, 2020. The RS VIII is located in the southeast of the province and encompasses 16 municipalities. The estimated population in 2010 was 1,150,290 inhabitants^16^.

The sampling frame was made up of the list of facilities from the Federal Registry of Health Care Facilities (REFES, by its acronym in Spanish) as of May 13, 2020. All state-funded health care facilities were included, with inpatient care whether it be general or specialized in maternal and child care, from the RS VIII municipalities. Santa Teresita Municipal Hospital was included, however, the Military Hospital from Tandil was excluded because the facility belongs to another administrative units. The list was comprised by 25 facilities which formed the First Stage Units. Municipal Health Secretaries from the RS VIII were requested to provide the total number of workers from those 25 facilities. A total of 8,617 workers conformed the Secondary Sampling Units.

In order to calculate the sample size, the population considered was the total number of workers from each facility. Regarding the proportion expected, the situation demanding the largest sample size was take into consideration (50%) With a confidence level of 95%, precision of 5% and design effect of 2, a sample size of 736 was obtained, but expanded to 1,000 participants in anticipation to a non-response rate possibility.

Two strata were considered: I) interregional hospitals (2 facilities) II) regional, subregional, municipal, local hospitals and health care units with inpatient care (23 facilities). The sample was proportionately distributed among the strata according to their size. All units from stratum I were included. The workers were selected using simple random sampling in each unit. In stratum II, the sampling was carried out using two-stage cluster sampling; 50% of the units were selected using probabilities proportionate to their size. The sample was distributed between them with a fixed fraction for all clusters (15%). In this last stage, the allocation of subjects to the sampling was made on the basis of the workers list provided by the selected facilities. The order number was assigned to the corresponding worker according to their position on the list ranked alphabetically.

All workers who signed an informed consent were included. Participants with contraindications for phlebotomy were excluded.

Data collection was carried out by means of a self-administered paper questionnaire with three sections: health care worker identification (ID), symptoms history, and clinical history. Previously validated questions were included, they were taken from the COVID-19 Suspicious Case Notification Form of the National Ministry of Health^17^ and from the risk assessment protocol for health care workers recommended by the WHO within the framework of early-response seroepidemiological investigations during the pandemic^10^. The recommendation to introduce an ethnic variable in the public health information system during COVID-19 was carried out in the pre-census joint design process with the National Institute of Statistics and Censuses (INDEC, by its acronym in Spanish) and the Network of Indigenous Professionals which was filed before the National Ministry of Health to promote the documentation of COVID-19 occurrences in indigenous populations^18^.

The variables and their categories were the following: Sex (male/female/other); Age (18–24, 25–34, 35–49, 50–64, 65 and older), Indigenous or Afro ancestry (yes/no); Occupation (orderly, nurse, pharmacist, surgical technologist, physical therapist, medical doctor, nutritionist, dentist, administrative/management personnel, admission/reception, cooking/catering, laboratory, cleaning, maintenance, security, laundry personnel, psychologist, radiologist, social worker, other); Weekly hours worked (number); In-site work during the Social, Preventive and Mandatory Isolation (ASPO, by its acronym in Spanish) (always, sometimes, never), Concurrent work (yes, in another health care facility; yes, somewhere else, but not in a health care facility, no); Use of personal protective equipment (PPE) (always, as recommended, most of the time, occasionally, never, I don’t know when to use it); History of close contact with a confirmed case of COVID-19 (yes, no, unknown); Presence of symptoms during the last two weeks (For each symptom: yes, no); Ongoing smoking habits (yes, no); Comorbidities (for each of them: yes, no); Pregnancy in progress (yes, no); Use of immunosuppressant medication (yes, no, unknown); History of sample taken in nasopharyngeal swabs by RT-PCR (yes, no, date, result).

A telephone hotline was made available so participants could contact the research team members in case they had inquiries when filling out the questionnaire. The laboratory personnel from each facility took blood samples from the participants. The serum or plasma samples (EDTA, citrate, heparin) were sent to the National Institute of Epidemiology “Dr. Juan H. Jara”, (INE by its acronym in Spanish) to be analyzed according to the applicable regulations^19^. Fasting for, at least, 3 hours was a requirement for the collection of the sample which was obtained by phlebotomy (5 ml) and collected in tubes previously labeled with the ID of each participant.

The test used was COVIDAR IgG co-developed by CONICET, Instituto Leloir, Universidad de San Martín y Laboratorio Lemos SRL authorized by ANMAT PM 1545–4. It consists of an immuno-enzymatic non-competitive heterogeneous assay based on an indirect in-vitro detection method for specific IgG antibodies for the SARS-CoV-2 spike protein in human serum or plasma samples^20^. The assay has a 100% specificity measured for pre-pandemic samples. The sensitivity determined in patients who had positive SARS-CoV-2 RT-PCR was 74% in the first 3 weeks from symptom onset and, at least, 91% in samples taken after 21 days from symptom onset. It is worth mentioning that in order to carry out studies where seroprevalence is low, it is necessary to use a high-specificity serological method.

In case isotype IgG antibodies were detected, IgM would be determined using the COVIDAR IgM test (ANMAT PM 1545-5). If IgM was detected, the collection of a nasopharyngeal and oropharyngeal swab sample was indicated on the worker so as to rule out viral activity using RT-PCR. In such situation, the worker would be isolated and close contacts would be actively traced.

A database was formulated using Epi Info^TM^ (version 7) in order to upload the questionnaires and serological results. A descriptive analysis was carried out on the basis of a distribution of qualitative variables in absolute and percentage values. The quantitative variables were summarized using measures of central tendency and dispersion. Tables and graphics were used to show the information.

The prevalence of infection was calculated for each stratum defined. The estimations were presented with a 95% confidence interval (CI). In all estimations, weighting was used. The total weight of each observation stemmed from multiplying the base weight by the non-response coefficient. The expanded sample consisted of 8,514 workers. In order to process the database, R 3.6.3 language, functions of tidyverse and epiR packages running in environment R Studio 1.2.5001 were used^21–23^.

The investigation initiative was approved by the INE Research Ethics Committee registered under code CE00264 in the National Registry of Health Research (RENIS, by its acronym in Spanish) authorized by the Central Ethics Committee no. 059/2019. Upon voluntary participation acceptance, each worker selected provided a signed informed consent.

## Results

Fieldwork took place between June 3 and July 6, 2020. The questionnaires and the informed consents were sent in enclosed envelops and distributed in each facility on the vehicles used to transport the samples used in RS VIII. A previously designated representative from each hospital was in charge of inviting the selected workers, informing them about the study objectives, and organizing the assignment of data collection.

The RS VIII administration contacted the municipal administration and health care departments of the facilities selected via videoconference, the study was presented, and operative concerns for implementation were established.

After the database clearance stage, 738 health care workers who complied with the blood test and filled in the questionnaire were included. The general response rate obtained was 73.80%. This number is nearly coincidental with the sample size calculated initially (736) that had been expanded to 1,000 participants in anticipation to a non-response rate possibility. Table 1 shows the response percentage per facility selected.

**Table 1:** Hospitals selected in the study and response rate. Sanitary Region VIII, Buenos Aires, Argentina, June 2020.

| Hospital (Municipality) | Sample obtained | Calculated sample | Response rate |
| --- | --- | --- | --- |
| Htal. M. Dr. "Arturo Illia" (Villa Gesell) | 48 | 48 | 100.00 |
| Htal. Local General de Coronel Vidal (Mar Chiquita) | 24 | 25 | 96.00 |
| Htal. de Niños Dr. "Débilio Blanco Villegas" (Tandil) | 16 | 17 | 94.12 |
| Htal. M. "Ana Rosa S. de Martínez Guerrero" (Madariaga) | 48 | 52 | 92.31 |
| Htal. Comunitario Dr. Dionisio José "Pepe" Olachea (Pinamar) | 49 | 54 | 90.74 |
| Htal. Subzonal M."Dr. Felipe A. Fosatti" (Balcarce) | 66 | 75 | 88.00 |
| Htal. M. "Dr. Carlos Macías" (La Costa) | 59 | 69 | 85.51 |
| Htal. M. "Gaspar M. Campos" (Lobería) | 26 | 32 | 81.25 |
| Htal. M. "Dr. Emilio Ferreyra" (Necochea) | 52 | 64 | 81.25 |
| Htal. M. de Santa Teresita (La Costa) | 41 | 51 | 80.39 |
| Htal. M."Dr. Marino Cassano" (General Alvarado) | 32 | 42 | 76.19 |
| Htal. Interzonal Especializado Materno Infantil "Don Victorio Tetamanti" (General Pueyrredón) | 141 | 190 | 74.21 |
| Htal. M. "Ramón Santamarina" (Tandil) | 56 | 96 | 58.33 |
| Htal. Interzonal General de Agudos "Dr. Oscar Alende" (General Pueyrredón) | 80 | 185 | 43.24 |

71.83% were female (N = 6,115, CI: 70.87–72.78), 3.00% were indigenous or afro descendants N = 255, CI: 2.65–3.38). The majority of occupations were included. Medical doctors and nurses represented more than half the amount of total health care workers. The health care workers worked in average 36.39 hours per week. 87.21% of the workers (N = 7,425, CI: 86.48–87.90) worked during the ASPO (Table 2). The average age was of 43.45 years old (19 youngest and 73 oldest). The age distribution showed a near-Standard distribution (Graph 1).

**Table 2:** Absolute and percentage distribution of socio-labour variables in health workers, Sanitary Region VIII, Buenos Aires, Argentina, June 2020 (N = 8,514).

| Variables | Frecuency | Percentage | CI 95% | Accumulated percentage |
| --- | --- | --- | --- | --- |
| **Sex** |  |  |  |  |
| Female | 6115 | 71.83 | 70.87-72.78 | 71.83 |
| Male | 2380 | 27.96 | 27.01-28.92 | 99.79 |
| Other | 10 | 0.12 | 0.06-0.22 | 99.91 |
| No data | 8 | 0.09 | 0.05-0.19 | 100.00 |
| **Age** |  |  |  |  |
| 18-24 | 138 | 1.62 | 1.37-1.91 | 1.62 |
| 25-34 | 1773 | 20.83 | 19.98-21.70 | 22.45 |
| 35-49 | 3949 | 46.39 | 45.33-47.45 | 68.84 |
| 50-64 | 2307 | 27.10 | 26.17-28.05 | 95.94 |
| 65 years old and above | 139 | 1.63 | 1.38-1.92 | 97.57 |
| No data | 207 | 2.43 | 2.13-2.78 | 100.00 |
| **Indigenous or Afro ancestry** |  |  |  |  |
| Yes | 255 | 3.00 | 2.65-3.38 | 3.00 |
| No | 8068 | 94.76 | 94.27-95.21 | 97.76 |
| No data | 191 | 2.24 | 1.91-2.66 | 100.00 |
| **Occupation** |  |  |  |  |
| Medical doctor | 2502 | 29.42 | 28.46-30.40 | 29.42 |
| Nurse | 1950 | 22.93 | 22.05-23.84 | 52.35 |
| Other | 826 | 9.71 | 9.10-10.36 | 62.06 |
| Administrative/management | 591 | 6.95 | 6.43-7.51 | 69.01 |
| Cleaning | 470 | 5.53 | 5.06-6.03 | 74.54 |
| Laboratory | 395 | 4.64 | 4.22-5.11 | 79.18 |
| Surgical technologist | 219 | 2.58 | 2.26-2.93 | 81.76 |
| Social worker | 168 | 1.98 | 1.70-2.29 | 83.74 |
| Admission/reception | 161 | 1.89 | 1.62-2.21 | 85.63 |
| Pharmacist | 149 | 1.75 | 1.49-2.05 | 87.38 |
| Security | 146 | 1.72 | 1.46-2.02 | 89.10 |
| Radiologist | 144 | 1.69 | 1.44-1.99 | 90.79 |
| Physical therapist | 121 | 1.42 | 1.19-1.70 | 92.21 |
| Cooking/catering | 119 | 1.40 | 1.17-1.67 | 93.61 |
| Laundry personnel | 113 | 1.33 | 1.11-1.60 | 94.94 |
| Psychologist | 100 | 1.18 | 0.97-1.43 | 96.12 |
| Maintenance | 89 | 1.05 | 0.85-1.29 | 97.17 |
| Orderly | 83 | 0.98 | 0.79-1.21 | 98.15 |
| Nutritionist | 80 | 0.94 | 0.76-1.17 | 99.09 |
| Dentist | 42 | 0.49 | 0.37-0.67 | 99.58 |
| No data | 36 | 0.42 | 0.27-0.67 | 100.00 |
| **Weekly hours worked** |  |  |  |  |
| Less than 30 hours | 1435 | 16.86 | 16.08-17.67 | 16.86 |
| 30-35 hours | 786 | 9.23 | 8.64-9.87 | 26.09 |
| 36-39 hours | 2938 | 34.51 | 33.51-35.53 | 60.60 |
| 40-47 hours | 1141 | 13.40 | 12.70-14.14 | 74.00 |
| 48 hours and above | 1961 | 23.04 | 22.15-23.94 | 97.04 |
| No data | 252 | 2.96 | 2.96-2.62 | 100.00 |
| **In-site work during ASPO** |  |  |  |  |
| Always | 7425 | 87.21 | 86.48-87.90 | 87.21 |
| Sometimes | 845 | 9.92 | 9.31-10.58 | 97.13 |
| Never | 198 | 2.33 | 2.03-2.67 | 99.46 |
| No data | 46 | 0.54 | 0.41-0.72 | 100.00 |

ASPO= Preventive and Mandatory Isolation (ASPO, by its acronym in Spanish)
CI= confidence interval

**Graph 1:**
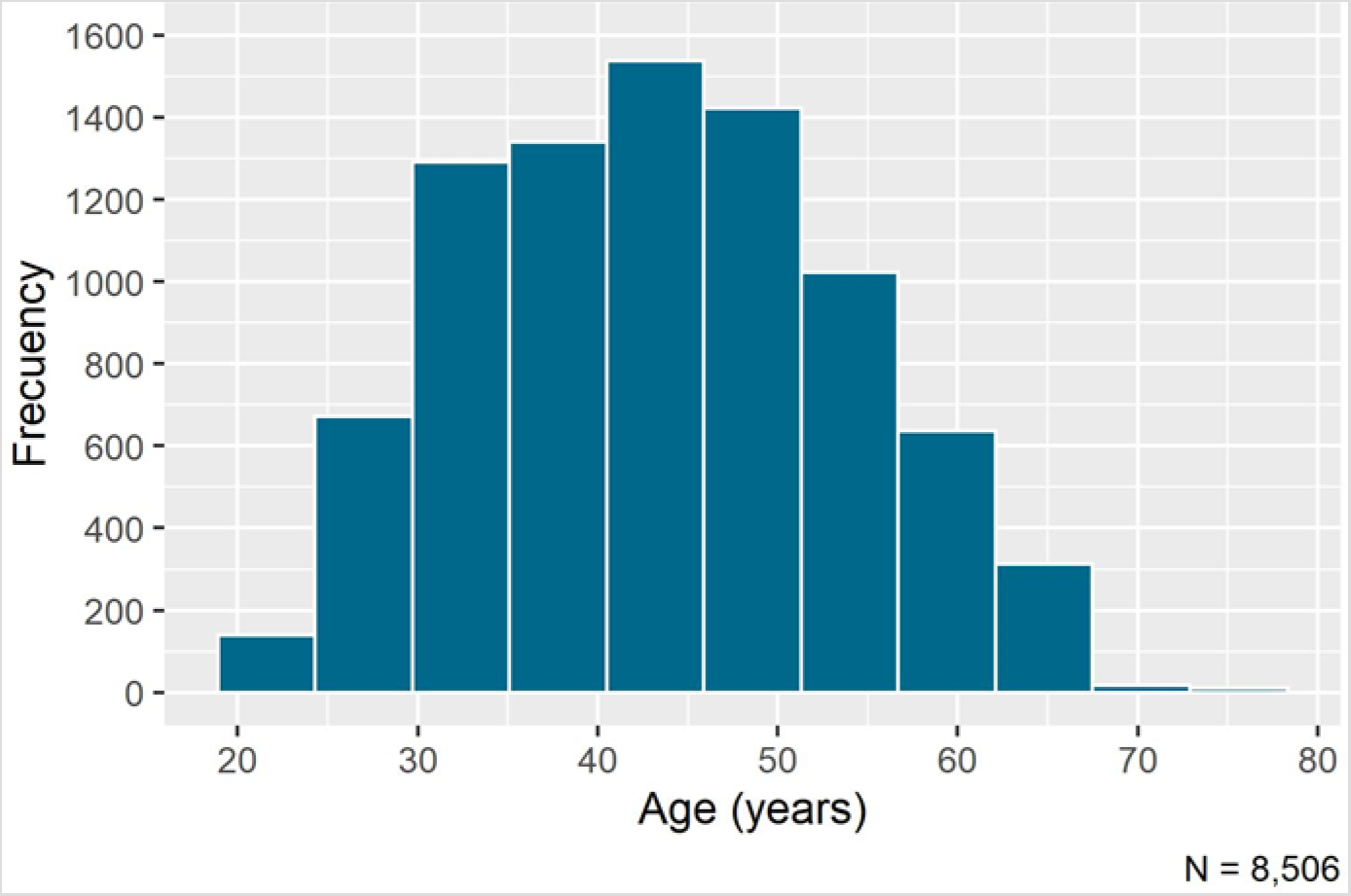
Age distribution of health workers, Sanitary Region VIII, Buenos Aires, Argentina, June 2020 (N = 8,514).

27.21 % (N = 2,317, CI: 26.28-28.17) worked in another health care facility during the ASPO apart from the hospital where they had been selected. 75.86% (N = 6,458, CI: 74.94–76.76) said they always wore PPE as recommended (Graph 2).

**Graph 2:**
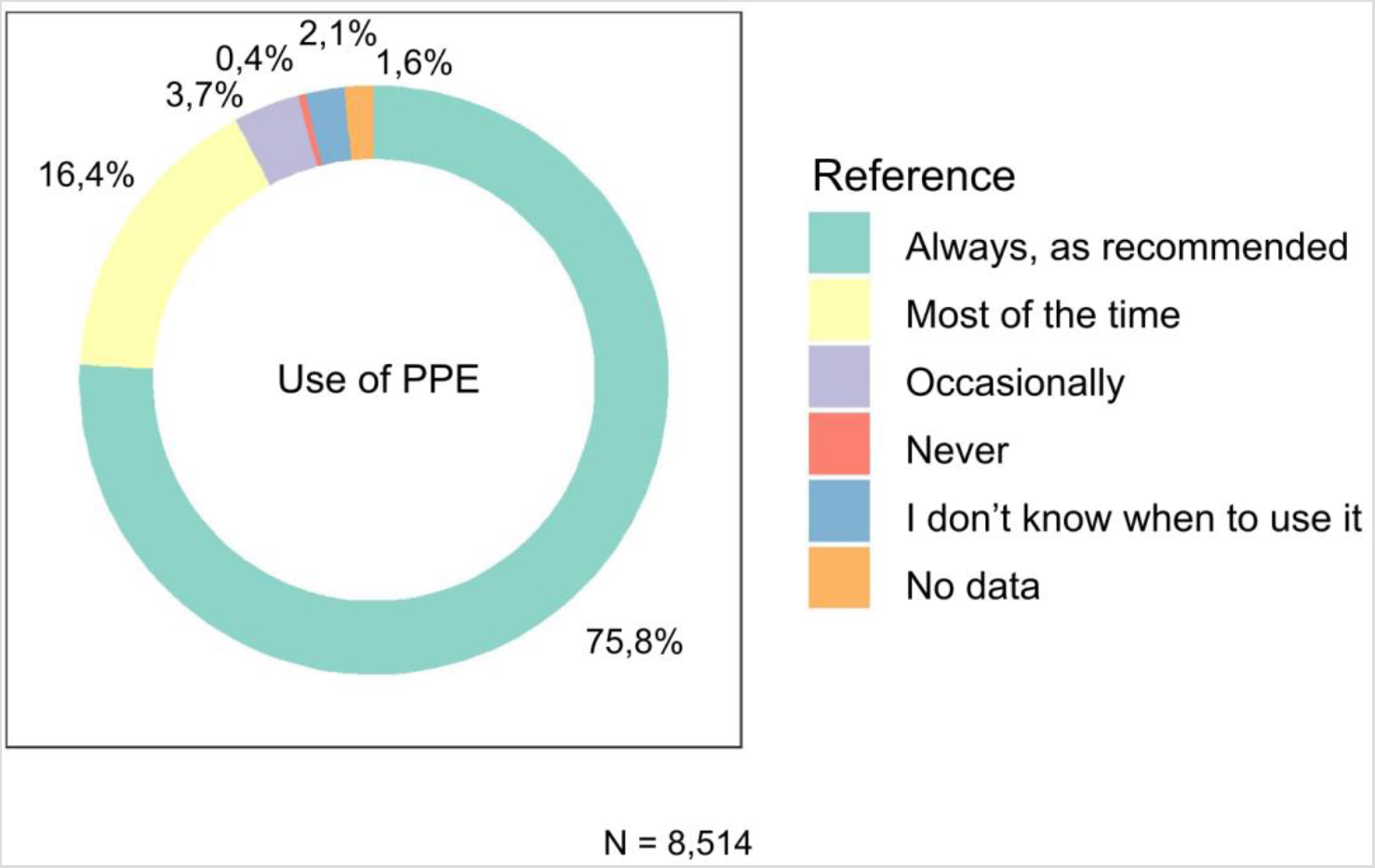
Frequency of use of personal protective equipment (PPE) by health personnel, Sanitary Region VIII, Buenos Aires, Argentina, June 2020 (N = 8,514).

Only 5.61% knew they had close contact with a COVID-19 confirmed case (N = 478, CI: 5.15-6.12) 4.60% of the workers had been subjected to a nasopharyngeal swab at some point in time (N = 392, CI: 4.18-5.07), and all tested negative.

Regarding the history of symptoms from the beginning of ASPO, headache was the most frequent (19.44%, n = 147), followed by runny nose 10.49 % (n = 76), less than 1% of the workers had fever (Table 3).

**Table 3:** Frequency of symptoms presentation (combined frequency) in health workers, Sanitary Region VIII, Buenos Aires, Argentina, June 2020.

| Symptoms | Frecuency* | Percentage |
| --- | --- | --- |
| Headache | 147 | 19.92 |
| Runny nose | 76 | 10.30 |
| Muscle aches | 66 | 8.94 |
| Cold | 48 | 6.50 |
| Cough | 42 | 5.69 |
| Fatigue | 35 | 4.74 |
| Abdominal pain | 25 | 3.39 |
| Diarrhoea | 22 | 2.98 |
| Agitation | 21 | 2.85 |
| Nausea / vomiting | 18 | 2.44 |
| Sore throat | 11 | 1.49 |
| Shortness of breath | 6 | 0.81 |
| Other respiratory symptoms | 6 | 0.81 |
| Loss of smell | 5 | 0.68 |
| Fever | 3 | 0.41 |
| Loss of taste | 2 | 0.27 |

High blood pressure was the most frequent comorbidity on 11.65% of the workers (n = 86); Diabetes mellitus was in second place (6.78%, n = 50) followed by asthma/COPD 6.10% (n = 45). 2.96% (N = 252, CI: 2.62-3.34) were receiving some kind of immunosuppressant treatment. Among the female, 0.36% were pregnant (N = 22, CI: 0.24–0.54). 22.54% smoked at the moment the survey was carried out (N = 1,919, IC: 21.66-23.44).

Five workers had positive SARS-CoV-2 IgG in epidemiological weeks 24 (2 cases), 25, 26, and 27, all had negative IgM. Of the cases two were medical doctors, one was a psychologist, one a nurse and one handled clothes (sewing). The average age of the cases was 35 years old(27 youngest and 46 oldest), four were female and one male. Only one of them did not work during the ASPO. One person worked in another health care facility apart from the selected hospital. They all informed to have always used PPE. None of them had knowledge of having been in close contact with a confirmed case of coronavirus. They did not have history of relevant comorbidities. Two of them were asymptomatic. The symptoms informed by the remaining three cases were: headache, diminished sense of smell and taste, muscle aches, cold and runny nose. None of the five workers with positive IgG had been subjected to a nasopharyngeal swab, and did not informed having been in close contact with a confirmed case of COVID-19.

The SARS-CoV-2 infection seroprevalence of health care workers in public hospitals with inpatient care whether it be general or specialized in maternal and child care of the RS VIII was 0.75% (CI: 0-8.13; standard error: 3.76). As per the strata, prevalence was 0.62% for interregional hospitals (CI: 0-9.82) and 0.82% (CI: 0-9.28) for regional, municipal, and local.

## Discussion

To the best of our knowledge, this was the first seroepidemiological study on health care workers carried out in Argentina during the COVID-19 pandemic with health care workers from the public subsector in the first line of response to the infection.

The prevalence detected was low, inferior to 1% with no significant difference between interregional hospitals and smaller ones. The findings of this investigation represent an important baseline to monitor the status of the pandemic. Four months after the emergence of the first cases in Argentina, less than one in one hundred health care workers of public hospitals have developed detectable antibodies for SARS-CoV-2. Aspects concerning the effectiveness and duration of the humoral immune response still remain to be determined, however, considering that the presence of IgG antibodies could be associated to a short-term protection, these findings call for attention to the fact that the majority of health care workers are still susceptible of being infected. This concern has been backed up by other seroprevalence studies carried out on the general public in countries that had even experienced a peak of the pandemic^14, 24^.

This investigation was carried out on health care workers in one of the 12 Sanitary Regions of the province with the largest number of inhabitants in Argentina. At the time of the study, there was scarce community transmission, which may have had an impact on the low prevalence found. The apparent low risk of infection is in harmony with the scarce amount of workers who informed history of close contact with a confirmed case of COVID-19.

In various countries around the world, the results of seroprevalence studies carried out during the COVID-19 pandemic on health care workers were dissimilar and inconsistent depending on the sampling strategy applied. It is interesting, however, that countries that experienced a rapid contagion curve with epidemics that collapsed the response capacity of the health care system have found relatively low prevalence results.

Besides, it is well-known that the use of PPE lowers the infection risks health care workers are exposed to^26–27^. In our study, a quarter of the workers informed not to have used it all the time. Such ratio should draw attention to the need to review the availability of PPE, training on its correct use, and investigation on aspects related to adherence to use.

Another worth-mentioning aspect is multiple occupations of the health care workers which may be associated to a work overload. Not an insignificant percentage informed they worked at another facility besides the facility selected for the study. Previous investigations to this pandemic highlighted the work overload as an infection risk factor on health care workers^28^. In addition, this matter should be considered at the moment of planning epidemiological surveillance and outbreak studies during the pandemic because of the possibility of spreading the infection on diverse health centers if proper use of PPE is not reinforced.

Even though the number of workers on whom antibodies were detected was low (five), two of them were asymptomatic. These findings, as well as the findings from other studies^29–30^ reinforce the need to harmonize passive epidemiological surveillance with serological surveillance on health care workers which may contribute to monitor the transmission dynamics and to evaluate infection control measures.

This study has strengths and weaknesses. Regarding its strengths, we can highlight the random sampling strategy which allowed to get a direct measure of the infection dimension on one of the most vulnerable population groups during the pandemic. There was a high overall response rate which clearly shows the commitment of the individuals involved and also contributed to obtain reliable estimations. As of May 1 of this year, methodological biases were found in a systematic review of the SARS-CoV-2 seroprevalence results, mostly due to omission of random sampling in their target population. On the other hand, at this moment, which is almost synchronous with the preparation stage of this investigation field, only 14 countries in the world had reported seroprevalence results as part of completed or ongoing studies^25^.

Regarding the weaknesses, the small amount of cases found did not allow to carry out an infection risk analysis according to different characteristics of the health care workers such as occupation or comorbidity history. The fieldwork done in order to obtain the required data for the sampling frame implied a great effort on the part of the research team since the information available was not integrated in only one database and had to be developed. In this regard, we highlight the need to improve the health care information systems in Argentina to allow for the possibility to access appropriate records.

## Conclusions

This investigation constitutes a baseline of the dimension of infection on health care workers in an area of Argentina with low viral transmission at the moment this work was carried out. The findings reinforce the importance of including this type of studies in the framework of active epidemiological surveillance strategies on vulnerable populations in order to monitor the infection tendency and the percentage of susceptible individuals.

## Data Availability

Data not available

## Acknowledgements

To the Hospital Directors included in the investigation and the representatives designated in each of them, for their cooperation during the fieldwork. To Leticia Gerbi and Agostina Quadrelli, from RS VIII, for their great effort in obtaining the workers list necessary to develop the sampling frame. To Hérctor Garcialoredo, for his work as data entry. To Marcelo Zotta, Silvina Lavayén, Lucía López Miranda, Verónica Poncet and Carlos Cimmino, from the Laboratory of Instituto Nacional de Epidemiología “Dr. Juan H Jara”. To COVIDAR Group for providing the ELISA kits. Without them, this investigation would not have been possible.

